# DNA methylation models of protein abundance across the lifecourse

**DOI:** 10.1101/2024.06.13.24308877

**Authors:** Scott Waterfield, Paul Yousefi, Matt Suderman

## Abstract

**Background:** Multiple studies have shown that DNA methylation (DNAm) models of protein abundance can be informative about exposure, phenotype and disease risk. Here we investigate the capacity of DNAm to capture non-genetic variation in protein abundance across the lifecourse.

**Methods:** We evaluated the performance of 14 previously published DNAm models of protein abundance (episcores) in peripheral blood from a large adult population using the Avon Longitudinal Study of Parents and Children (ALSPAC) at ages 7-24 and their mothers antenatally and in middle age (N range = 145-1464). Episcore-protein associations were evaluated with and without adjustment for genetics. New age-specific episcores were trained in ALSPAC and evaluated at different ages.

**Results:** Of the 14 Gadd episcores, 10 generated estimates associated with abundance in middle age, 9 at age 24, and none at age 9. Eight of these episcores explained variation beyond genotype in adulthood (6 at age 24; 7 at midlife). At age 9, the abundances of 22 proteins could be modelled by DNAm, 7 beyond genotype of which one trained model generated informative estimates at ages 24 and in middle age. At age 24, 31 proteins could be modelled by DNAm, 19 beyond genotype, of which 5 trained models generated informative estimates at age 9 and 8 in middle age. In middle age, 23 proteins could be modelled, 13 beyond genotype, of which 3 were informative at age 9 and 7 at age 24.

**Conclusions:** We observed that episcores performed better at older ages than in children with several episcores capturing non-genetic variation at all ages.

## Introduction

In humans, there are over 20,000 protein coding genes, many of which have well-characterised patterns of tissue-specific expression and function^1^. Aberrant protein expression is known to play important roles in the development of many diseases. Changes in protein abundance allow for the diagnosis and prognosis of many diseases^2^ and may be influenced by a variety of biological and environmental causes including genotype^3^ and gene regulators such as DNA methylation (DNAm)^4^.

DNAm is a reversible epigenetic modification which is linked to both genetic and environmental factors and is known to regulate gene expression^5^. In mammals this is primarily carried out by adding a methyl group to a cytosine residue which is followed by a guanine residue (termed CpG site). Recently, models have been developed using DNAm levels measured at several CpG sites to predict protein abundance. For example, the Grimage model^6^ includes DNAm models of a number of plasma proteins which together are capable of accurately predicting an individual’s time to death. It is notable that this model which uses DNAm to proxy proteins performs better as an indicator of time to death than models derived directly from DNAm. This has been hypothesised to be due to the protein models capturing functionally relevant DNAm variation, which may be missed when directly regressing 100,000s of individual CpG sites onto time to death directly.

Motivated by the success of the Grimage model, Gadd et al^7^ carried out a large-scale analysis of the viability of DNAm to index protein abundance and found this to be the case for 109 of 953 proteins. These models, called episcores, were projected into an external sample and associations were found with a number of morbidities. Both the model training and evaluation of their associations with morbidity were carried out in older adult populations, as such it is not clear whether the performance of these models transfers to younger populations, especially children.

An analogous investigation of genetic indices of protein abundance has also been carried out by Xu et al^8^, which generated models called polygenic scores (PGS) for 825 of 2692 proteins. Given that genotype is established at fertilisation and is largely stable throughout life whereas disease processes and exposures tend to differ by life stage, DNAm models of protein abundance will likely capture additional variation associated with disease progression and environmental exposures beyond that of genetic models alone. However, since DNAm variation is at least partially (∼20%) determined by genotype^9^, we expect that DNAm models of protein abundance may capture some of the variation captured by genetic models.

Here, we ask how episcores trained in later adulthood perform across the lifecourse and to what extent DNAm may capture age-specific variation in protein abundance. We first test if Gadd episcores trained in older adult populations transfer to younger populations in ALSPAC (ages 9 and 24) and if they replicate in similarly aged populations in ALSPAC ( mothers of study participants at middle age). Next, we consider if episcores projected into DNAm at one time point are capable of predicting protein expression at a future time point. Following this, we determine to what extent episcores capture variance in protein beyond genotype. Finally, we train episcores for 88 proteins quantified in ALSPAC children at ages 9 and 24 and in their mothers at middle age (referred to as ALSPAC episcores), evaluate their transferability between time points, and determine to what extent these episcores capture variance in protein levels beyond the genotype (see Figure 1 for general overview of analyses).].

**Figure 1.**
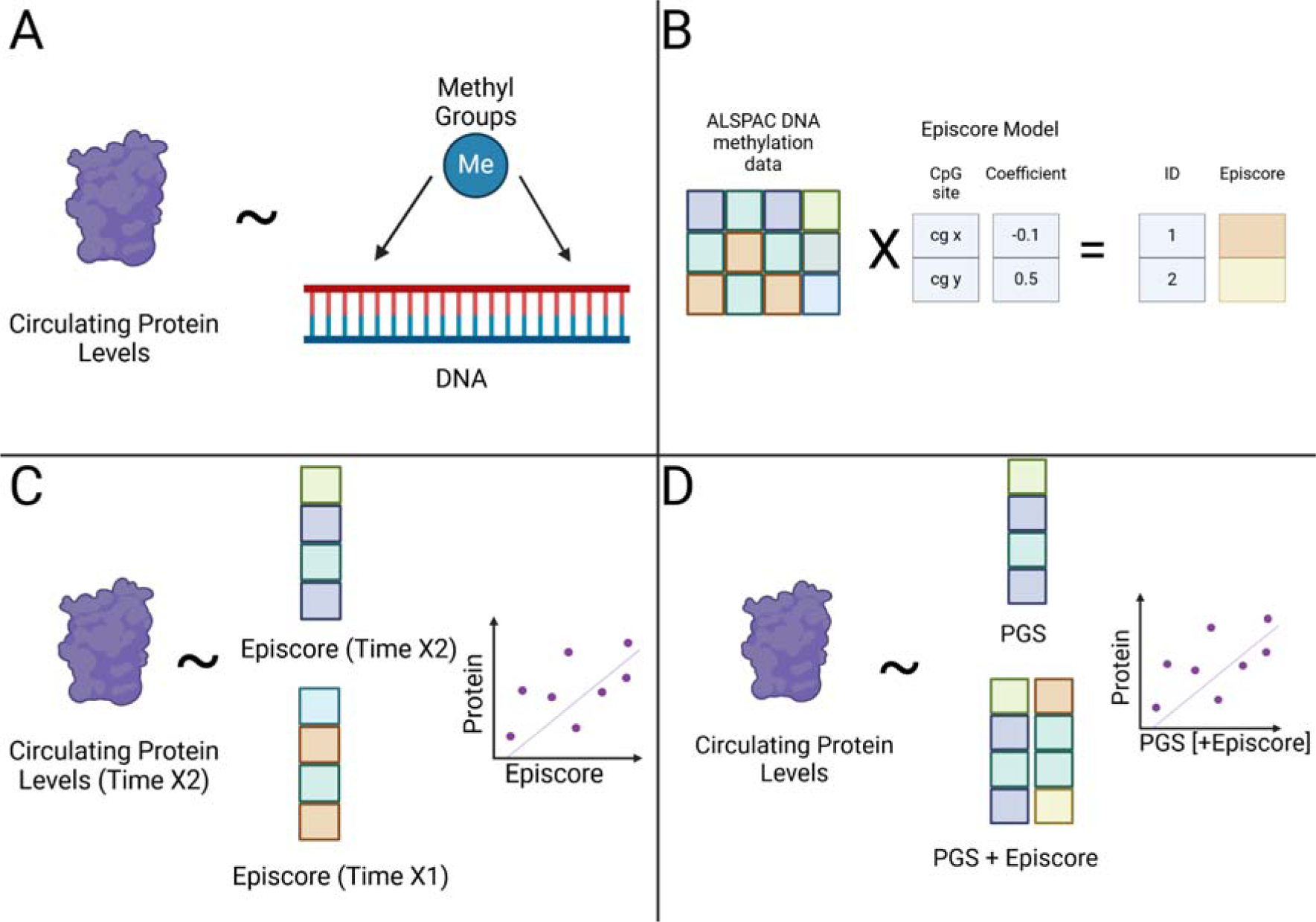
General overview of methods and analyses. (A) Process of training episcores. (B) Projecting episcores into ALSPAC. (C) Cross-sectional and prospective prediction of protein levels using episcores. (D) Incorporating genetic models to determine non-genetic information capture of episcores. Created with BioRender.com

## Methods

### Study population

ALSPAC is a prospective birth cohort study in south west England. Pregnant women resident in one of the three Bristol-based health districts with an expected delivery date between April 1, 1991 and December 31, 1992 were invited to participate. The study has been described elsewhere in detail and ethical approval for the study was obtained from the ALSPAC Ethics and Law Committee and the Local Research Ethics Committees^10–12^.

ALSPAC initially enrolled a cohort of 14,541 pregnancies, from which 14,062 live births occurred, with 13,998 alive at 1 year. Follow-up has included parent and child completed questionnaires, links to routine data and clinic attendance.

Research clinics were held when these offspring participants were approximately 9, 10, 11, 13, 15, 18, and 25 years old. Data for 25 years of age were collected and managed using REDCap electronic data capture tools hosted at the University of Bristol. REDCap (Research Electronic Data Capture) is a secure, web-based software platform designed to support data capture for research studies^13^. The study website contains details of all the data that is available through a fully searchable data dictionary http://www.bristol.ac.uk/alspac/researchers/access/.

### DNA methylation measurements

As part of the ARIES project (http://www.ariesepigenomics.org.uk), a sub-sample of ALSPAC mother–child pairs had DNA methylation measured using the either the Illumina Infinium HumanMethylation450 Beadchip (450K array) or the Illumina Infinium MethylationEPIC BeadChip (EPIC array). In blood samples collected from ALSPAC children, DNAm was measured at ages 7, 9, 15-17 and 24 and from the mothers during the study child pregnancy (‘antenatal’) and approximately 18 years later (‘midlife’). All DNA methylation wet-lab and preprocessing analyses were performed at the University of Bristol as part of the ARIES project and has been described in detail previously^14^. The subsamples used in our study are described in Table 1.

**Table 1.**
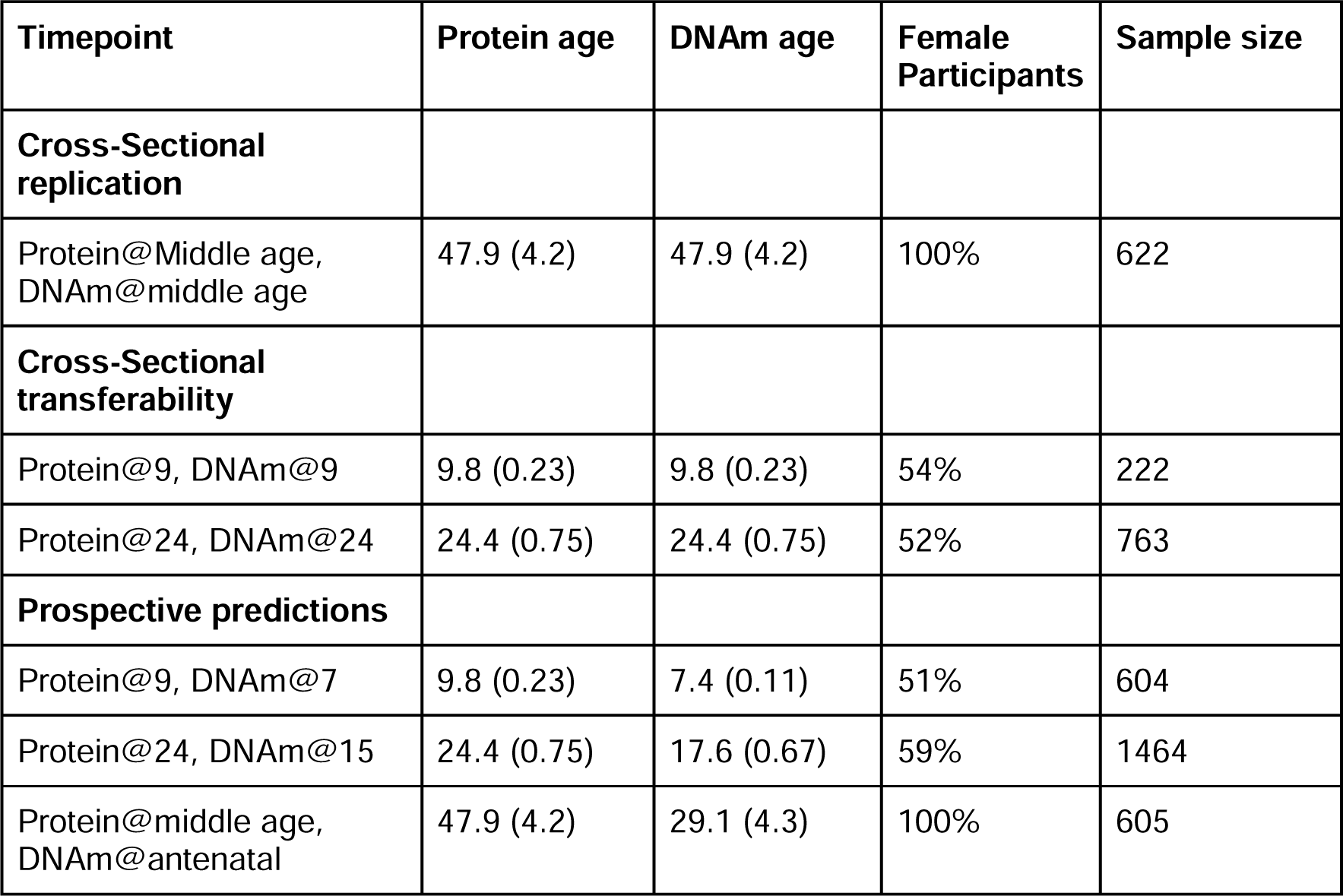
Sample sizes for comparisons of DNAm estimates of protein abundance and direct Olink measurements in ALSPAC. age is measured in mean years (standard deviation (SD)).

### Proteomic measurements

Protein measurements were previously carried out for a total of 9000 blood plasma samples in ALSPAC using the Olink Target 96 inflammatory panel, split between non-fasted children at age 9 (3005), fasted children at age 24 individuals (3027) and 2968 fasted mothers at midlife. This data has been previously described in full by Goulding et al^15^. Efforts were made to maximise the number of children with protein measurements at both 9 and 24 and the number of mother-child pairs with protein measurements.

### Previously published DNAm models of protein abundance (Gadd episcores)

Recently published^7^ DNAm models for abundance of 109 proteins (Gadd episcores) were trained using DNAm and protein abundance measured in blood samples from the KORA study^16^. Of these 109 proteins, the abundances of 12 were measured in ALSPAC using the Olink platform. These corresponded to 14 Gadd episcores (2 proteins had both proteins measured by both Somascan and Olink platforms) which were then projected into ALSPAC DNAm. These episcores were projected into ALSPAC at ages 7, 9, 15 and 24 in the ALSPAC children, and at antenatal and middle age timepoints in the mothers (see Table 1).

### Genetic models of protein abundance

Genetic models of protein abundance for 308 Olink- and 2,348 Somascan-measured proteins were published by Xu et al.^8^. Proteins with measurements on both platforms have two PGS, one for each platform. For these, only the PGS with the strongest correlation with measured protein abundance was used for further analysis.

### Novel DNAm models of protein abundance in ALSPAC (ALSPAC episcores)

We trained de novo linear DNAm models of protein abundance in ALSPAC participants at three life stages (age 9, age 24, in middle age). Elastic Net as implemented by the *glmnet* R package^17^ was used to train models for all 88 measured Olink proteins with all autosomal CpGs sites measured on the EPIC array as possible features. Protein values were residualised by sex prior to training. Model hyperparameters, alpha and lambda, were jointly optimized by 5-fold cross validation. Coefficients for ALSPAC-trained episcores for all three timepoints are available in Supplementary File 1.

Proteins with with mean cross-validated R >0.10 in ALSPAC at a specific time point, were modelled using data from all participants with data at that time point and then projected into DNAm measured at other time points with matched protein abundance measurements. For example, episcores trained in middle age maternal participants were also projected into DNAm measured at ages 9 and 24. DNAm in ALSPAC was measured using a mix of 450k and EPIC arrays. In cases where models would be trained and projected into DNAm data generated using a mix of arrays, models trained on the subset of CpG sites common to both arrays.

### Evaluating episcore performance across the life course

Gadd episcores were assessed for replication by projecting them into DNAm measured in ALSPAC middle age mothers, which is demographically similar to the datasets used by Gadd to build these scores. Replication was deemed successful if model estimated and measured protein abundances were correlated at Pearson correlation coefficient threshold R > 0.10 and P < 0.05. We also assessed whether Gadd episcore performance was transferable to less similar data by projecting them into DNAm measured in ALSPAC offspring at age 9 and 24, again using a R > 0.10 and, P < 0.05 threshold.

The predictive capacity of Gadd episcores was evaluated by projecting the episcores into ALSPAC at age 7 and age 15 and evaluating their correlation with protein measurements at ages 9 and 24, respectively.

The capacity of DNAm to model protein abundance at a given time point in ALSPAC was assessed within a cross-validation framework (5 folds). DNAm was considered capable of modelling a protein if the mean correlation between estimates and measurements across folds was at least 0.1. Models were then trained for such proteins using all available data at that time point. Transferability of these models to other time points was assessed by projecting these models into DNAm other time points and calculating correlations between model estimates and protein measurements. A model was considered transferable for correlation R > 0.10 and P < 0.05.

### Comparison of episcore and genetic measure of protein abundance

Capture of non-genetic protein variation was evaluated through the use of F-tests to compare two linear models, the first including just a published PGS to predict protein abundance and a second including both an episcore and the PGS to predict protein abundance (threshold for reporting: F-test P < 0.05). This was carried out at each cross-sectional time point (age 9, age 24, middle age). For the ALSPAC episcores, F-tests were performed within a cross-validation framework (5 folds) so that episcores could be trained and evaluated within the same dataset.

### Effect of protein stability and variance on model performance

Correlations of protein levels between ages 9 and 24 were calculated to determine the longitudinal stability of protein abundance. The relationship between protein stability and episcore transferability was evaluated by calculating the correlations between the protein stability correlations and the ALSPAC model transferability correlations from age 9 to 24, and vice versa.

## Results

### Replication and transferability of Gadd episcores in ALSPAC

Gadd episcores were projected into DNAm measured in ALSPAC at middle age to assess replication. Concurrent Olink measurements for 12 proteins matched 14 Gadd episcores since there are 2 episcores (trained from Somascan and Olink measurements) for both of CXCL11 and CXCL10. Of these, 10 episcores replicated (P < 0.05 and Pearson’s R > 0.1, Figure 2 and Table 2).

**Figure 2.**
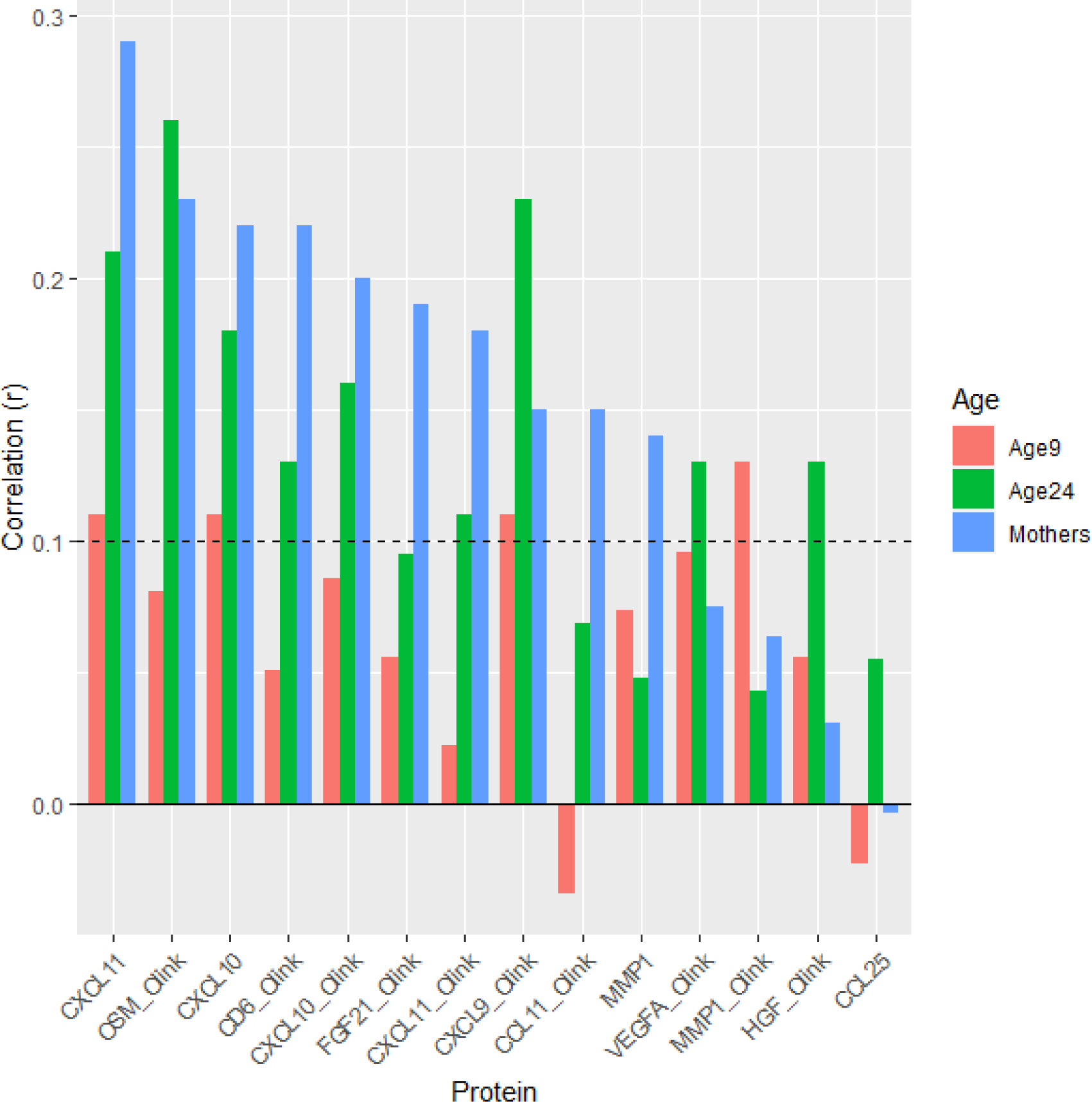
Replication and transference of Gadd episcores in ALSPAC at ages 9 (N = 222), 24 (N = 763), and at midlife (N = 622).

**Table 2.** Replication and transferability of Gadd episcores in ALSPAC at ages 9 (n = 222) and 24 (n = 763) and midlife (N = 622). Correlations above a nominal threshold (R > 0.10, P < 0.05) are highlighted in bold.

|  | age 9 |  | age 24 |  | Middle age |  |
| --- | --- | --- | --- | --- | --- | --- |
| Episcore | R | P | R | P | R | P |
| CXCL11_Soma | 0.11 | 0.11 | **0.21** | **4.8e-09** | **0.29** | **2.2e-13** |
| OSM_Olink | 0.081 | 0.23 | **0.26** | **6.6e-13** | **0.23** | **6e-09** |
| CXCL10_Soma | 0.11 | 0.11 | **0.18** | **1e-06** | **0.22** | **1.6e-08** |
| CD6_Olink | 0.051 | 0.45 | **0.13** | **0.00029** | **0.22** | **5.9e-08** |
| CXCL10_Olink | 0.086 | 0.2 | **0.16** | **6.5e-06** | **0.2** | **4.8e-07** |
| FGF21_Olink | 0.056 | 0.4 | 0.095 | 0.0087 | **0.19** | **2.6e-06** |
| CXCL11_Olink | 0.022 | 0.74 | **0.11** | **0.0036** | **0.18** | **4.6e-06** |
| CXCL9_Olink | 0.11 | 0.11 | **0.23** | **2e-10** | **0.15** | **0.00013** |
| CCL11_Olink | -0.034 | 0.62 | 0.069 | 0.057 | **0.15** | **0.00024** |
| MMP1_Soma | 0.074 | 0.27 | 0.048 | 0.18 | **0.14** | **0.00057** |
| VEGFA_Olink | 0.096 | 0.15 | **0.13** | **0.00021** | 0.075 | 0.06 |
| MMP1_Olink | 0.13 | 0.055 | 0.043 | 0.23 | 0.064 | 0.11 |
| HGF_Olink | 0.056 | 0.4 | **0.13** | **0.00047** | 0.031 | 0.43 |
| CCL25_Soma | -0.023 | 0.73 | 0.055 | 0.13 | -0.0034 | 0.93 |

We also projected Gadd episcores into DNAm measurements from ALSPAC offspring at ages 9 and 24 with concurrently available Olink protein measurements to evaluate whether they were transferable to younger populations. Here, only four episcores transferred to age 9 (Pearson’s R > 0.1 but none at P < 0.05) and nine to age 24 (P < 0.05 and Pearson’s R > 0.1, Figure 2 and Table 2).

### Gadd episcores have predictive capacity for protein levels

To determine the predictive capacity of Gadd episcores, we assessed whether Gadd episcores projected into DNAm measured at one time point could predict protein abundance measured at a later time point (Figure 3). No episcores projected into age 7 DNAm were predictive of protein abundance at age 9 (Supplementary Table 1). Two episcores at age 15 were predictive of protein abundance at age 24 (Supplementary Table 2). Seven episcores projected into DNAm measured during pregnancy predicted protein abundance at middle age, approximately 18 years later (Supplementary Table 3).

**Figure 3.**
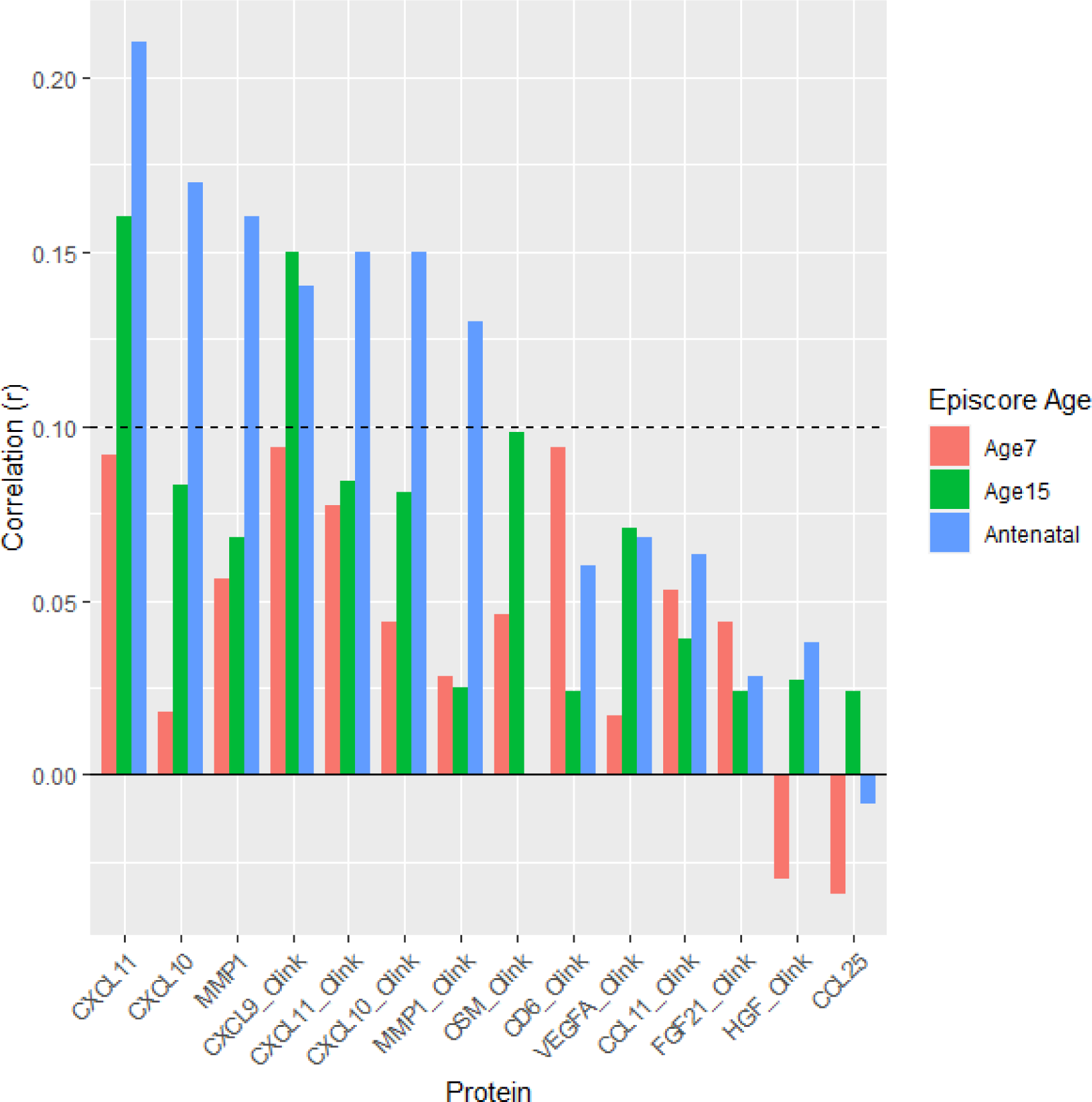
Prediction of future protein levels in ALSPAC by Gadd episcores from age 7 (DNAm) to 9 (protein abundance) (N = 604), age 15 to 24 (N = 1464), and pregnancy to middle age 18 years later (N = 605).

### Gadd episcores explain protein variance beyond genotype

Next we asked whether Gadd episcores explained variance in protein abundance beyond polygenic scores (PGS). PGS, Gadd episcores, and protein abundance data were available for eight proteins in ALSPAC at ages 9, 24 and middle age. We evaluated the additive explanatory value of Gadd episcores by performing F-tests to compare a baseline model with just a PGS predicting protein abundance to a model additionally including the corresponding Gadd episcore (Methods).

At age 9, no Gadd episcores showed evidence (F-test P < 0.05) of explanatory beyond the PGS alone (Supplementary Table 4).However, at age 24, Gadd episcores for five proteins explained protein variance beyond the PGS: CD6, CXCL10, CXCL11, OSM, VEGFA (Figure 4A,Supplementary Table 5). For two proteins, CXCL11 and OSM, Gadd episcores explained more than 5% of the variance in protein abundance beyond polygenic scores.

**Figure 4.**
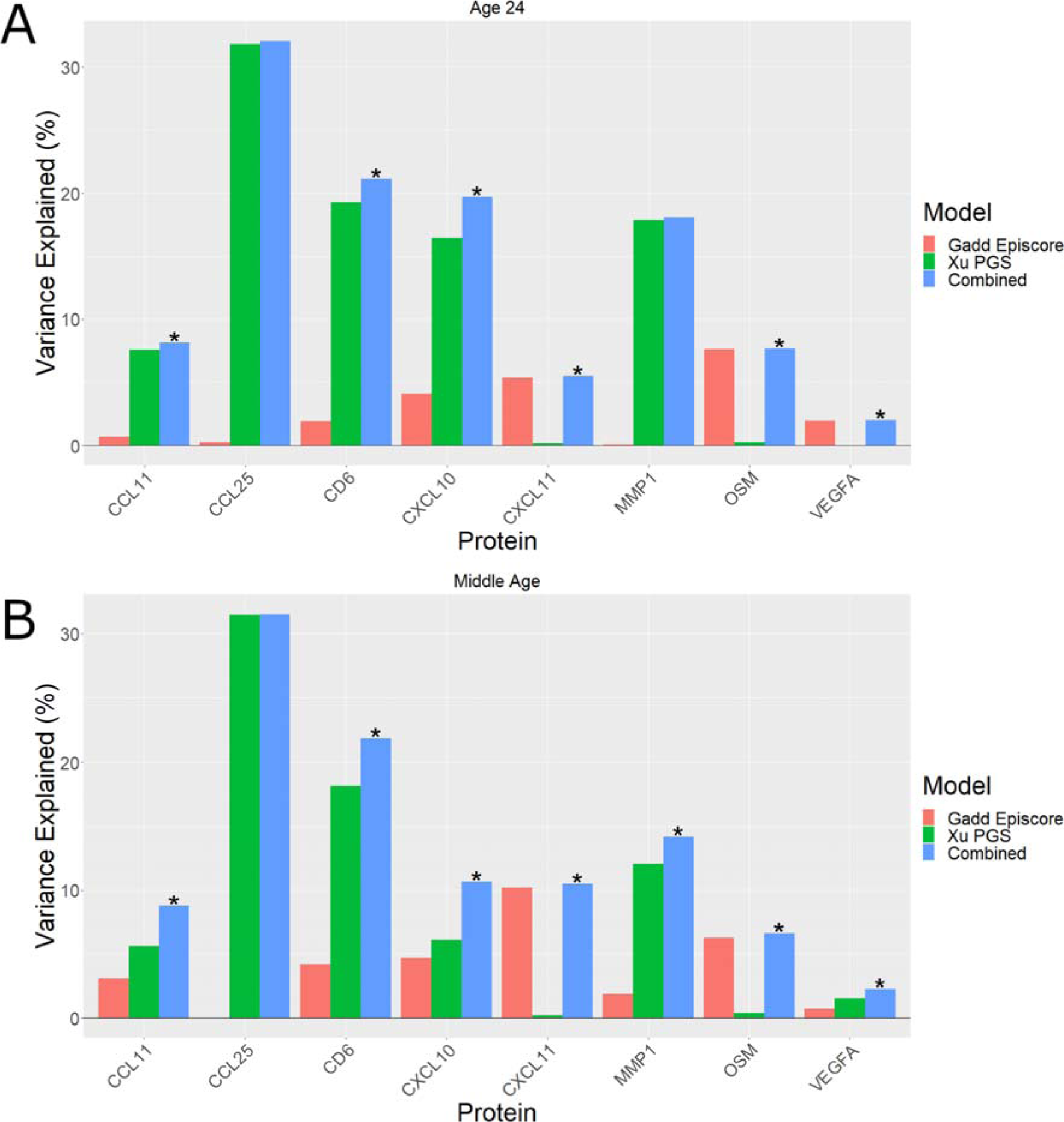
Variance in protein abundance explained by Gadd episcores and PGS separately and combined in ALSPAC at (A) 24 years, and (B) middle age. Combined models which explain significantly more variance than the genetic model alone are marked with an asterisk (**\***) (F-test P < 0.05).

Similarly, in middle aged mothers, Gadd episcores captured protein variance beyond the genotype for seven proteins: CCL11, CD6, CXCL10, CXCL11, MMP1, OSM, VEGFA (Figure 4B,Supplementary Table 6). Here again, the Gadd episcores for CXCL11 and OSM explained more than 5% of variance in protein abundance beyond polygenic scores.

### ALSPAC-trained episcores transfer across ages

To evaluate the transferability of episcores trained within ALSPAC, we trained episcores using the matched DNAm and protein abundance data available in ALSPAC at ages 9 and 24 and in middle age. For each time point, we used cross-validation (CV) to evaluate model performance and evaluated transferability of models for proteins with sufficiently strong CV performance (mean CV R > 0.1). We observed strong performance for 22, 32 and 23 of the 88 proteins at ages 9, 24 and middle age, respectively.

We then trained episcores for these proteins using all available data at each given time point and asked if they transferred to the other time points. Of the 22 episcores trained at age 9, only IL18R1 transferred to older ages, in this case to both age 24 and middle age (Figure 5A). For the 31 episcores trained at age 24, five transferred to age 9, and eight transferred to middle age (Figure 5B). Finally, of the 23 episcores trained in middle age, three transferred to age 9 and seven transferred to age 24 (Figure 5C; Supplementary Tables 7-9).

**Figure 5.**
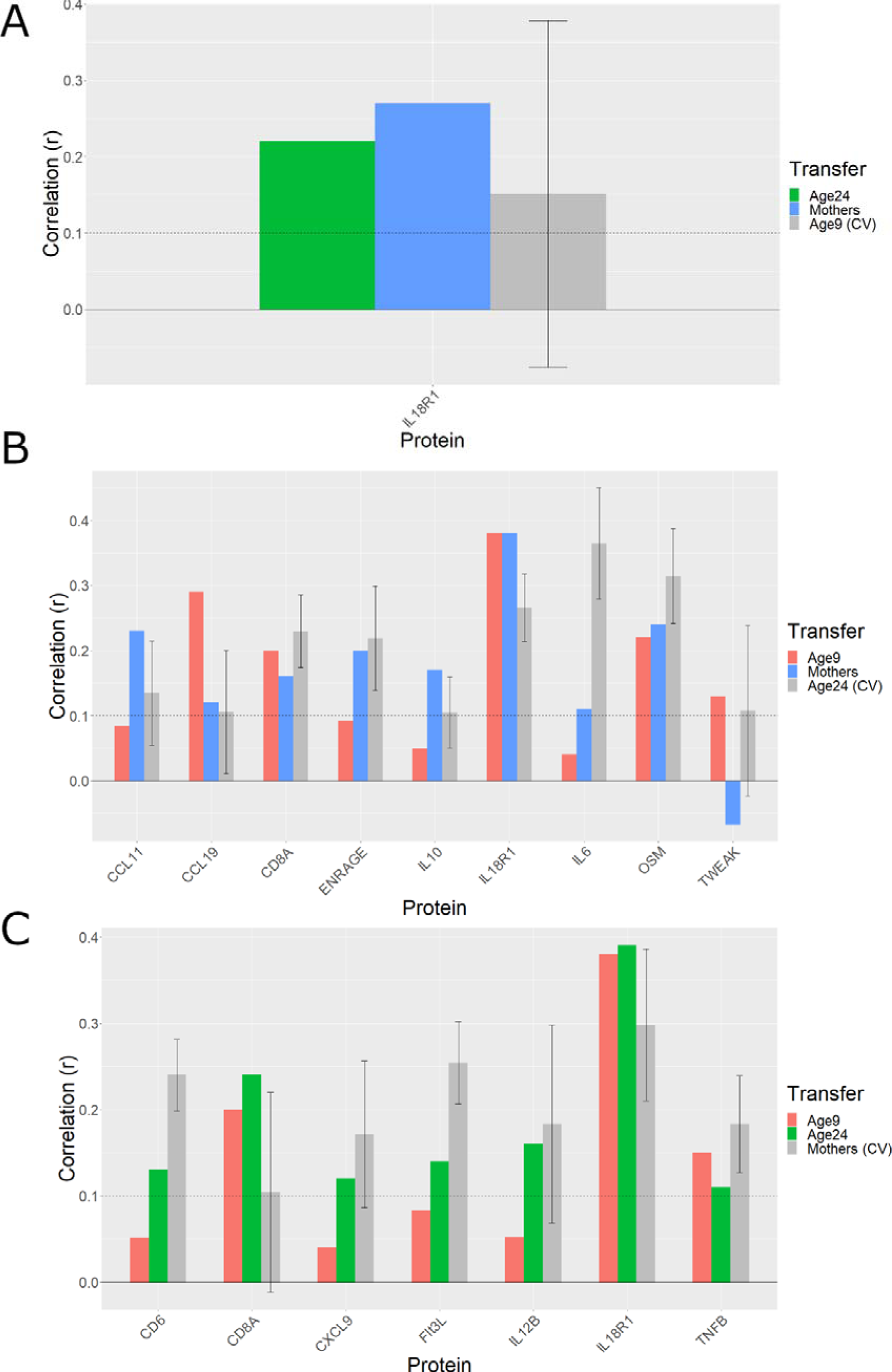
Transferability of episcores trained in ALSPAC. (A) Episcore trained at age 9 and transferred to age 24 (N = 319-763) and middle age (N = 274-622). (B) Episcores trained at age 24, transferred to ages 9 (N range = 145-336) or to middle age (N range = 274-622). (C) Episcores trained at middle age and transferred to ages 9 (N = 145-336) or 24 (N = 319-763). Error bars represent +/- 1 standard deviation (SD) for CV performance within training time point.

Given that protein abundance may differ by sex, we asked whether transferability may differ by sex. All participants at middle age were women, so sex-specificity was evaluated between ages 9 and 24. Sex-specificity was evaluated by calculating sex-specific transfer correlations and then asking if they differed between the sexes (t-test). We observed no sex differences for the eight episcores that transferred into age 9 (t = 0.74, P =0.52) nor for the seven that transferred into age 24 (t = −0.01, P 0.99).

### Episcores trained in childhood and in adulthood explain non-genetic variation

For our ALSPAC-trained episcores, we again asked whether episcores explain variation in protein abundance beyond genotype. This was evaluated through the use of cross-validation to maximise power (Methods).

At age 9, seven of 12 episcores with a corresponding PGS explained protein variance beyond the PGS (F-test P < 0.05; Figure 6A, Supplementary Table 10). One episcore (*TGFalpha*) explained more than 5% of the variance in protein abundance beyond the PGS. At age 24, nineteen of the 21 episcores with a corresponding PGS explained protein variance beyond PGS (Figure 6B,Supplementary Table 11). Episcores for four proteins (CXCL11, IL6, OSM, TNFSF14) explained more than 5% of the variance in protein abundance beyond the PGS. In middle aged mothers, thirteen of 13 episcores with a corresponding PGS explained protein variance beyond the PGS (Figure 6C,Supplementary Table 12). Episcores for four proteins (CD6, CXCL11, Flt3L, OSM) explained more than 5% of variance in protein abundance beyond the PGS.

**Figure 6.**
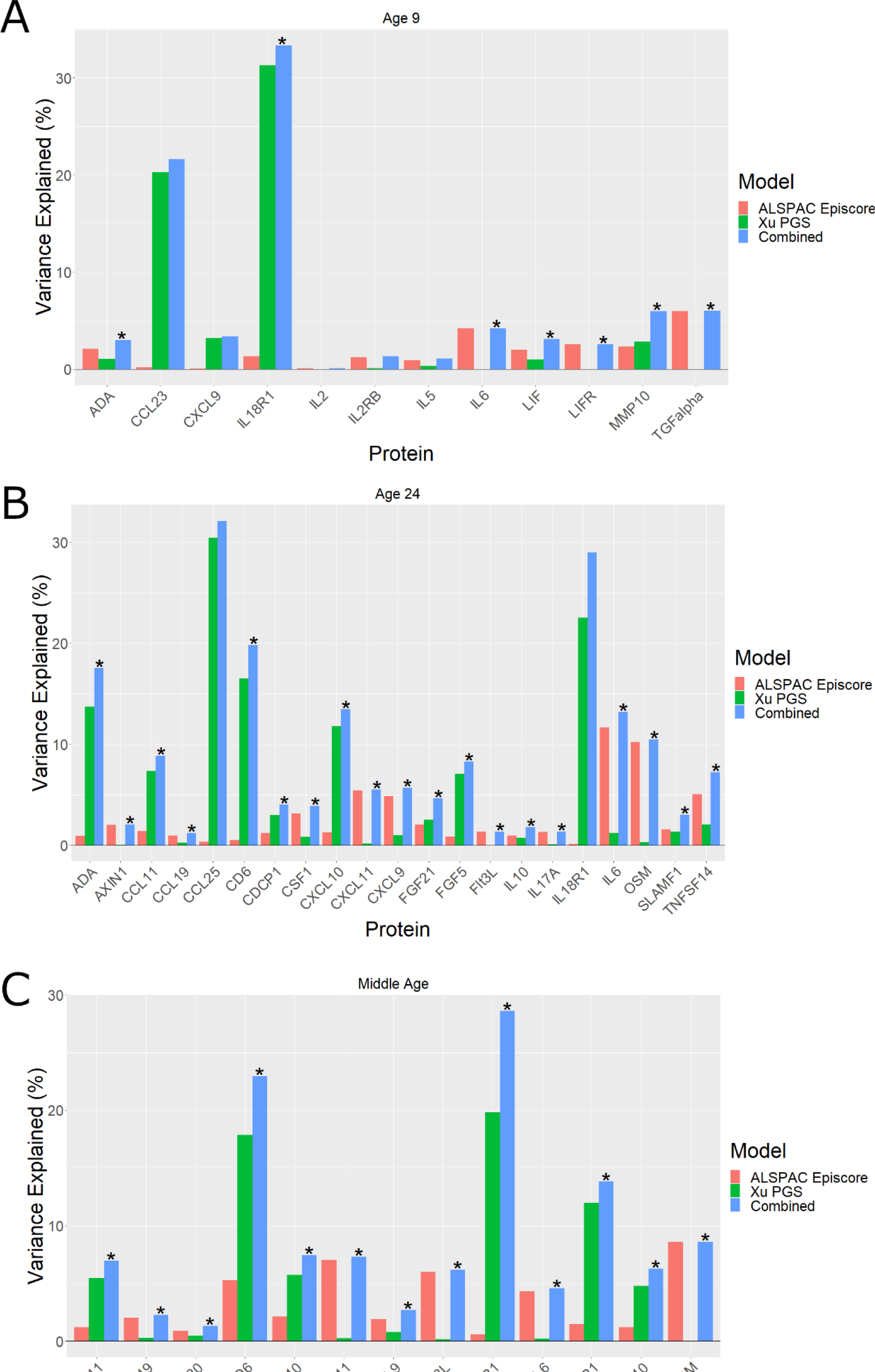
Variance in protein abundance explained by ALSPAC episcores and polygenic scores separately and combined at (A) age 9 (N range = 197-309), (B) 24 years (N range = 663-704), and (C) middle age (N range = 547-553). Combined models which explain more variance than the polygenetic score alone are marked with an asterisk (**\***) (F-test P < 0.05).

### Longitudinal protein stability doesn’t determine episcore transferability

To determine if the stability of protein levels affects transferability, we compare the longitudinal stability of proteins from age 9 to age 24 against the transferability DNAm models from age 9 to 24, and from age 24 to age 9. We observed weak positive correlations both from age 9 to 24 (R = 0.31, P = 0.15) (Figure 7A) and from age 24 to 9 (R = 0.23, P = 0.21) (Figure 7B).

**Figure 7.**
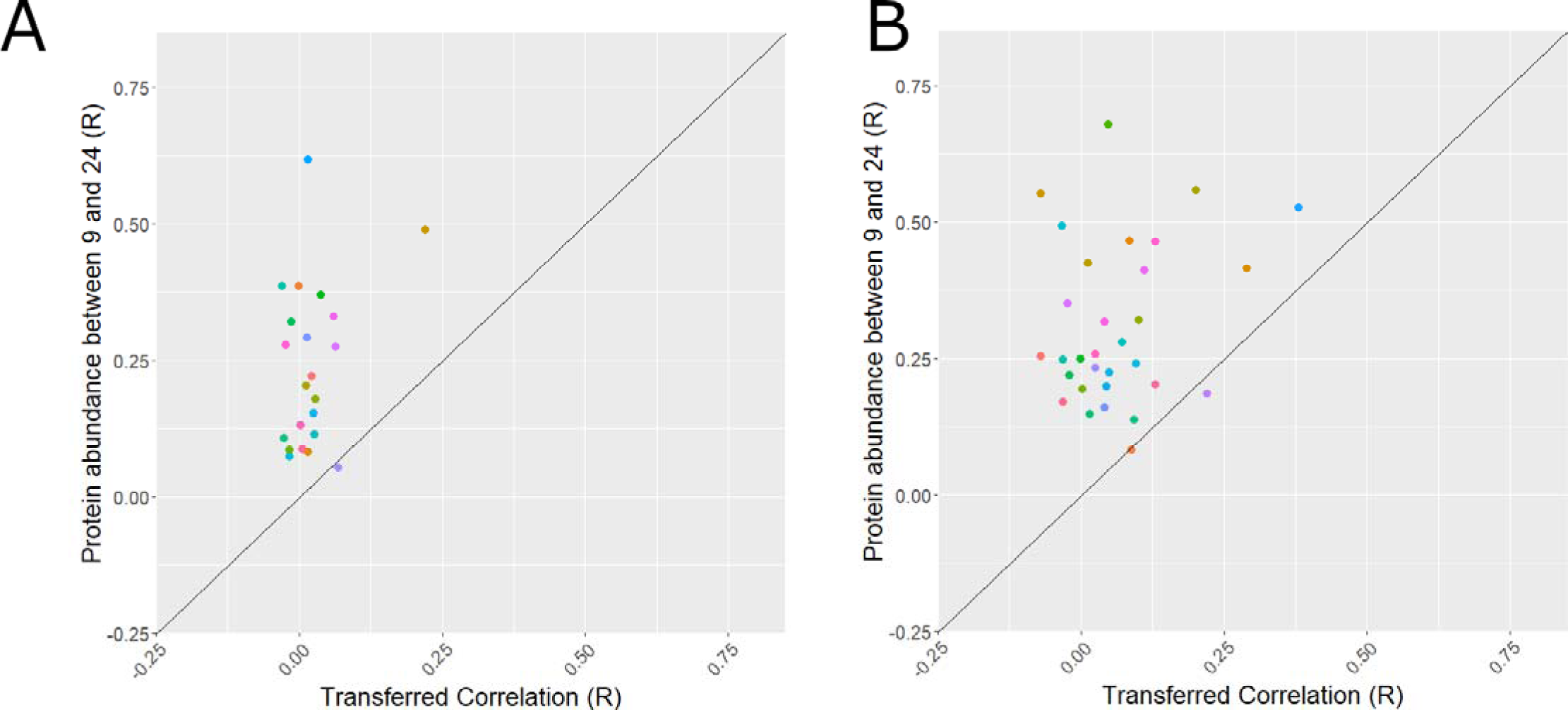
The association between longitudinal protein stability and ALSPAC episcore transferability. (A) Associations for transfers from age 9 models into age 24, (B) Associations for transfers from age 24 models into age 9.

## Discussion

Although Gadd et al.^7^ showed that DNAm can be used to estimate protein abundance, their episcores have never been tested in younger populations. Here, we have evaluated these episcores in ALSPAC participants at three different ages. We find that most of the Gadd episcores capture variance in protein abundance in young (age 24) and middle aged adults but very little in children (age 9).

We also examined the extent to which episcores capture non-genetic variation. We did indeed observe evidence for this for several proteins in young and middle age adults, whether the episcore was trained within or outside ALSPAC. We observed some but weaker evidence for this at age 9, but only for episcores trained at age 9. This is consistent with our understanding that protein abundance like DNA methylation is determined by a combination of both genetic and environmental factors, and suggests that molecular prediction of protein abundance should, where possible, integrate both genetic and epigenetic sources of information. We note here a potential bias of this analysis, as the Gadd episcores were only released should they pass a certain threshold in test data, whilst the Xu polygenic models were released for every protein tested. As such it is unlikely that the proportion of episcores shown to be successful over genetic models is truly representative. Training set sizes also differed quite significantly for episcore (n ∼ 500-1000 for Gadd episcores and n ∼ 250-750 for ALSPAC episcores) and polygenic models (n ∼ 3000-5000). Larger sizes for episcore training would almost certainly have increased the number of proteins with trained episcores and the proportion of protein variance captured.

To further examine the capacity of DNAm to capture protein abundance across the lifecourse, we trained new episcores in ALSPAC at ages 9 and 24, and in middle age. We find that the capacity of DNAm to capture protein abundance at age 9 is low compared to its capacity in adulthood at age 24 and in middle age. Transferability to and from age 9 was similarly poor and evidence for capture of protein variance beyond genotype weaker than for older ages. By contrast, we found the capacity of DNAm to capture protein variation at age 24 to be quite similar to middle age, including capacity to capture variation beyond genotype. In fact, this capacity was comparable to that of the Gadd episcores, also trained in mainly middle-age adults. This is somewhat unexpected given that Gadd et al. regressed out genetic variation from protein abundances prior to training episcores^7^, whilst we do not. Future work should determine if this step produces better models both for capturing protein variation and for health-related phenotypic variation ore generally.

The limitations of our study include small and highly variable sample sizes (N range = 145-1464), which introduces concerns around statistical power and comparability of results. The use of longitudinal data from the same familial cohort for use in model testing may have inflated model metrics (due to relatedness between mothers and children, and due to autocorrelation between time points for children with measurements in more than one time point). The Xu polygenic models were also trained in adults, so they may perform less well in children and young adults. Our conclusions are limited to the relatively small set of inflammatory proteins we examined in ALSPAC (88 proteins from the Olink inflammatory panel). Finally, we note that all episcore and PGS models discussed were trained using data obtained from white European populations, so our findings may not apply outside of this demographic.

## Authors’ Contributions

SW planned and carried out all analyses and wrote the first draft. MS and PY supervised the project and critically reviewed the manuscript.

## Data Availability

Data is available at request from https://www.bristol.ac.uk/alspac/researchers/access/

## References

1. Kim, M.-S. et al. A draft map of the human proteome. Nature 509, 575–581 (2014).

2. Aronson, J. K. & Ferner, R. E. Biomarkers-A General Review. Curr. Protoc. Pharmacol. 76, 9.23.1–9.23.17 (2017).

3. Melzer, D. et al. A genome-wide association study identifies protein quantitative trait loci (pQTLs). PLoS Genet. 4, e1000072 (2008).

4. Jones, P. A. Functions of DNA methylation: islands, start sites, gene bodies and beyond. Nat. Rev. Genet. 13, 484–492 (2012).

5. Attwood, J. T., Yung, R. L. & Richardson, B. C. DNA methylation and the regulation of gene transcription. Cell. Mol. Life Sci. 59, 241–257 (2002).

6. Lu, A. T. et al. DNA methylation GrimAge strongly predicts lifespan and healthspan. Aging 11, 303–327 (2019).

7. Gadd, D. A. et al. Epigenetic scores for the circulating proteome as tools for disease prediction. Elife 11, (2022).

8. Xu, Y., Ritchie, S. C., Liang, Y., Timmers, P. & Pietzner, M. An atlas of genetic scores to predict multi-omic traits. bioRxiv (2022).

9. Min, J. L. et al. Genomic and phenotypic insights from an atlas of genetic effects on DNA methylation. Nat. Genet. 53, 1311–1321 (2021).

10. Fraser, A. et al. Cohort Profile: the Avon Longitudinal Study of Parents and Children: ALSPAC mothers cohort. Int. J. Epidemiol. 42, 97–110 (2013).

11. Boyd, A. et al. Cohort profile: the ‘children of the 90s’—the index offspring of the Avon Longitudinal Study of Parents and Children. Int. J. Epidemiol. 42, 111–127 (2013).

12. Northstone, K. et al. The Avon Longitudinal Study of Parents and Children (ALSPAC): an update on the enrolled sample of index children in 2019. Wellcome Open Res 4, 51 (2019).

13. Harris, P. A. et al. Research electronic data capture (REDCap)--a metadata-driven methodology and workflow process for providing translational research informatics support. J. Biomed. Inform. 42, 377–381 (2009).

14. Relton, C. L. et al. Data Resource Profile: Accessible Resource for Integrated Epigenomic Studies (ARIES). Int. J. Epidemiol. 44, 1181–1190 (2015).

15. Goulding, N. et al. Inflammation proteomics datasets in the ALSPAC cohort. Wellcome Open Res. 7, 277 (2022).

16. Holle, R., Happich, M., Löwel, H., Wichmann, H. E. & MONICA/KORA Study Group. KORA--a research platform for population based health research. Gesundheitswesen 67 Suppl 1, S19–25 (2005).

17. Hastie, Qian & Tay. An Introduction to glmnet. CRAN R Repositary.

